# AI-based discovery of functional boundaries in the human brain from intraoperative electrophysiology

**DOI:** 10.64898/2026.05.02.26352297

**Authors:** Simon Leszek, Matthew R. Baker, Bryan T. Klassen, Michael A. Jensen, Gaby Ojeda Valencia, Klaus-Robert Müller, Kai J. Miller

**Affiliations:** Machine Learning Group, Technische Universität Berlin; Department of Neurosurgery, Mayo Clinic.; Department of Neurology, Mayo Clinic.; Department of Biomedical Engineering and Physiology, Mayo Clinic.; BIFOLD – Berlin Institute for the Foundations of Learning and Data; Department of Artificial Intelligence, Korea University.; Max Planck Institute for Informatics, Saarbrücken

**Keywords:** Deep Brain Stimulation, Microelectrode Recording, Thalamus, Artificial Intelligence

## Abstract

Neurosurgical and neuromodulation therapies such as deep brain stimulation (DBS) require millimeter-level accuracy to effectively target functional brain regions. Yet, many neuroanatomical boundaries remain invisible to current imaging and electrophysiology methods, limiting precision and contributing to suboptimal patient outcomes. Here, we introduce a self-supervised artificial intelligence (AI) framework that learns to delineate functional subregions directly from the spectral content of intraoperative local field potential (LFP) recordings, without the need for predefined biomarkers or anatomical labels. The framework identifies physiologic structure across the full spectrum of the signal and, through explainable AI (XAI), reveals the specific frequency components underlying these distinctions. Validated in the subthalamic nucleus (STN), the model aligned with clinically defined borders and rediscovered known beta oscillations. Applied to the motor thalamus in tremor patients, it consistently identified functional transitions corresponding to the ventral oralis posterior (Vop) and ventral intermediate (Vim) nuclei—regions where conventional methods fail to provide reliable boundaries. To assess clinical relevance, physiologically defined clusters were functionally evaluated using monopolar review data at their first DBS clinic postoperative visit, demonstrating distinct stimulation-response profiles across clusters and linking electrophysiologic segmentation to clinically meaningful programming outcomes. These findings demonstrate that intraoperative LFP recordings can be transformed into both a real-time guidance resource and a data-driven platform for biomarker discovery, establishing a foundation for more precise, individualized neuromodulation therapies and advancing our understanding of functional brain organization.

## Introduction

The success of surgical and neuromodulation therapies for brain disorders depends on millimeter-level accuracy in targeting functional neural circuits. For deep brain stimulation (DBS), even small deviations in electrode placement can compromise efficacy, cause side effects, require surgical revision, or prolonged stimulation programming [1–3]. This exemplifies a broader challenge in neurosurgery and biomedical engineering: Despite major advances in imaging and electrophysiology, generalizable intraoperative methods to define functional boundaries in the human brain remain limited. Addressing this gap is key for advancing neurological therapies and our broader understanding of human brain function.

For DBS lead placement surgery, clinicians have traditionally relied on visible imaging landmarks and physiological biomarkers for guidance [4, 5]. This approach is effective for certain structures such as the subthalamic nucleus (STN), where anatomical borders can be visualized with magnetic resonance imaging (MRI) and single-unit physiology on microelectrode recordings [3]. In contrast, other borders like the basal ganglia (ventral oralis posterior; Vop) and cerebellar (ventral intermediate nucleus; Vim) inputs to the motor thalamus are not detectable by standard imaging and electrophysiology approaches [6–9], making Vim DBS lead placement for medication-resistant tremor particularly challenging (Fig. 1A-B). Beyond single-unit physiology, local field potentials (LFPs) capture circuit-level dynamics and have yielded robust biomarkers in certain regions (i.e. beta oscillations in the STN; [10, 11]). However, their broader utility has been constrained by the reliance on predefined frequency bands and the difficulty of translating these signals into real-time surgical guidance.

**Fig. 1.**
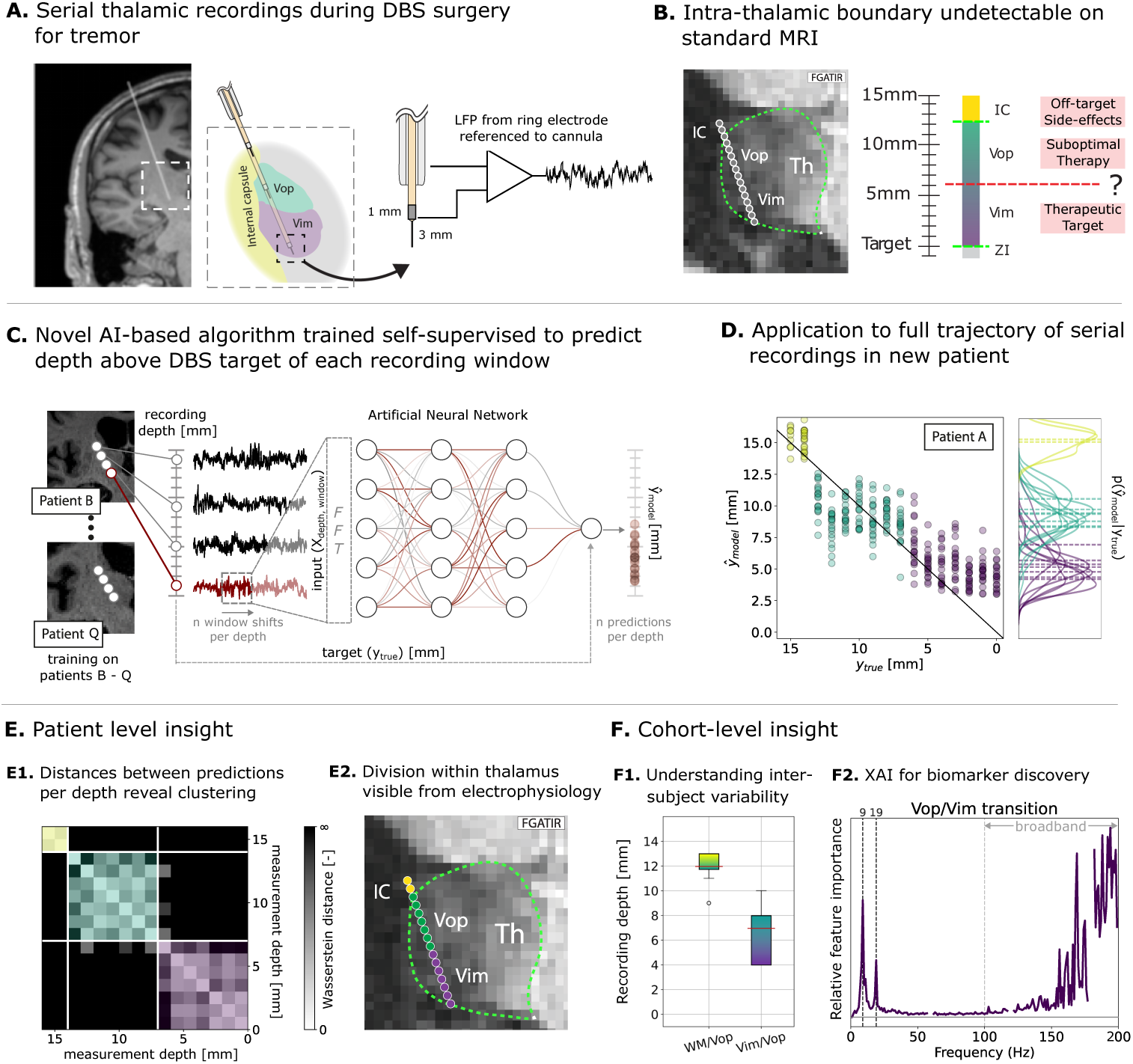
Overview on challenge of targeting within the motor thalamus (A-B), our proposed ThalaMAP framework (C-D), and the respective insights (E-F): **(A)** Thalamic recordings during DBS surgery for tremor are collected from a ring contact on the microelectrode. **(B)** Up to 15 serial depth recordings (at ∼1mm intervals, each lasting 30s) are made per DBS track on the way to the target (Vim sub-region). While the borders of the thalamus are visible on MRI, the internal motor thalamus border is not. **(C)** We split the recordings of each patient (leave-one-out) into 1 second windows and train an ANN to predict the respective recording depth in a self-supervised manner. **(D)** When applied to individual data traces from a new patient to predict depth, we find clusters which correspond to functional boundaries along the measurement trajectory. The clusters of patient A are visualized in **(E1)** and plotted back onto imaging anatomy alongside the clinically annotated transition of white matter and thalamus **(E2).** Patient specific mapping allows for a better understanding of inter-subject variability of functional regions **(F1)** and the application of eXplainable AI (XAI) enables the identification of (previously unknown) biomarkers **(F2).** Vim = ventral intermediate nucleus of the thalamus; Vop = ventral oralis posterior; Th = thalamus; IC = internal capsule.

To address this challenge, we developed a self-supervised artificial intelligence (AI)-based framework that learns directly from the spectral content of intraoperative recordings without predefined biomarkers or anatomical labels. Unlike conventional approaches that presuppose which frequency bands are relevant, our method identifies physiologic structure across the full spectral domain, revealing functional transitions that are otherwise undetectable. With the addition of explainable AI (XAI) methods, the framework identifies the specific frequency ranges that drive these distinctions.

We demonstrate the framework using LFPs from DBS microelectrode recordings (MER), first validating it in the STN, where it aligned with clinically defined borders and rediscovered known beta oscillations. We then applied it to the motor thalamus, where conventional imaging and physiology cannot reliably distinguish adjacent sub-regions, and the framework consistently revealed functional transitions corresponding to the Vop and Vim nuclei. Using explainable AI, we further identified several low frequency oscillations and broadband features, providing new insight into the physiologic signatures of motor thalamus organization. To assess clinical relevance, physiologically defined clusters were subsequently evaluated using postoperative monopolar review, demonstrating distinct stimulation-response profiles and linking electrophysiologic segmentation to clinical programming outcomes. In addition, we performed an intraoperative feasibility demonstration in which the framework was deployed in real time during DBS surgery, illustrating its potential for live neurosurgical integration. Together, these results show that intraoperative recordings can be transformed into both a guidance resource for surgical targeting and a discovery platform for new biomarkers, establishing a paradigm for individualized brain mapping with broad relevance across neurosurgery, neuroscience, and biomedical engineering.

## Results

### ThalaMAP: a framework for functional boundary discovery

To identify functional brain boundaries in the absence of robust imaging or physiological biomarkers, we use machine learning techniques from self-supervised and representation learning [12]. Inspired by early adoption of self-supervision in images [13], where spatial image patch information is employed, we propose modeling the MER recording distance (depth in mm) from the planned surgical target as a proxy task for self-supervision. This allows the model to incorporate information inherent to the expected order of transitioned regions along the measurement trajectory. For instance, a 15mm MER span targeting the Vim, typically starts within in the internal capsule (IC) then transitions into the Vop and ends in the Vim (*depth*_IC_ *> depth*_Vop_ *> depth*_Vim_). Consequently, we formulate this inference problem as a regression task, where the model predicts the recording depth using power spectral features from a sliding window across the raw LFP signal (Fig. 1C). We ensure generalization across patients with a leave-one-patient-out training strategy and guide the model to focus on the most relevant spectral features through model regularization (for details, see Materials and Methods).

Interestingly, the model’s depth predictions form consistent clusters across patients, revealing shared physiologic structure. Specifically, we observe high similarity in model predictions for recording points within each cluster, while similarity markedly decreases between clusters. The effect is especially notable at transition points between clusters, where prediction similarity drops sharply between adjacent depths, despite their physical proximity (Fig. 1D-E1). We conclude that the model has learned to map the signal’s spectral signature to the respective brain (sub-) region, which can be projected back onto MRI images in-plane with the recording trajectory [14] revealing previously invisible functional boundaries (Fig. 1E2). To gain insights into specific frequency signatures of different regions, we utilize methods from the field of explainable AI (XAI) [15–17] (Fig. 1F2), enabling unbiased, data-driven biomarker discovery. For further technical details see Materials and Methods.

### Validation on subthalamic nucleus

To validate our framework in a setting with a known ground truth, we applied ThalaMAP to a cohort of 8 patients receiving STN DBS for Parkinson’s Disease [3]. In this context, transitions from the white matter into the STN are both visible on MRI, and have robust single-unit and spectral (i.e. *β*-oscillations) biomarkers for direct targeting [3, 10, 11]. As described above, the AI model was trained in a leave-one-out fashion and predicted depth for the left out patient.

For STN patient A, model depth predictions form two distinct clusters, one for the top ten and one for the bottom three recording sites, consistent with clinically determined boundaries indicated by the dashed pink line (Fig. 2A). In figure 2B, the identified clusters and visible boundaries are plotted back onto MRI, showing perfect alignment. To determine physiological agreement we calculated the average PSD for each resulting cluster, showing clear *β* oscillation (12-30Hz) and broadband differences (1/f shape; Fig. 2C). For patients B-H we consistently found the same STN cluster at the bottom of the trajectory with sub-millimeter precision to clinical boundaries (limited by 1mm macroelectrode size). Cohort-level XAI relevance scores (relative importance of frequency *i* for the model output) highlighted particular importance of *β* oscillations, as expected. These findings not only show highly accurate model assignment to clinically-determined STN borders, they also validate that the model has learned an electrophysiologically meaningful strategy, demonstrating its generalization potential for novel biomarker discovery. Detailed results per patient are presented in Appendix A.

**Fig. 2.**
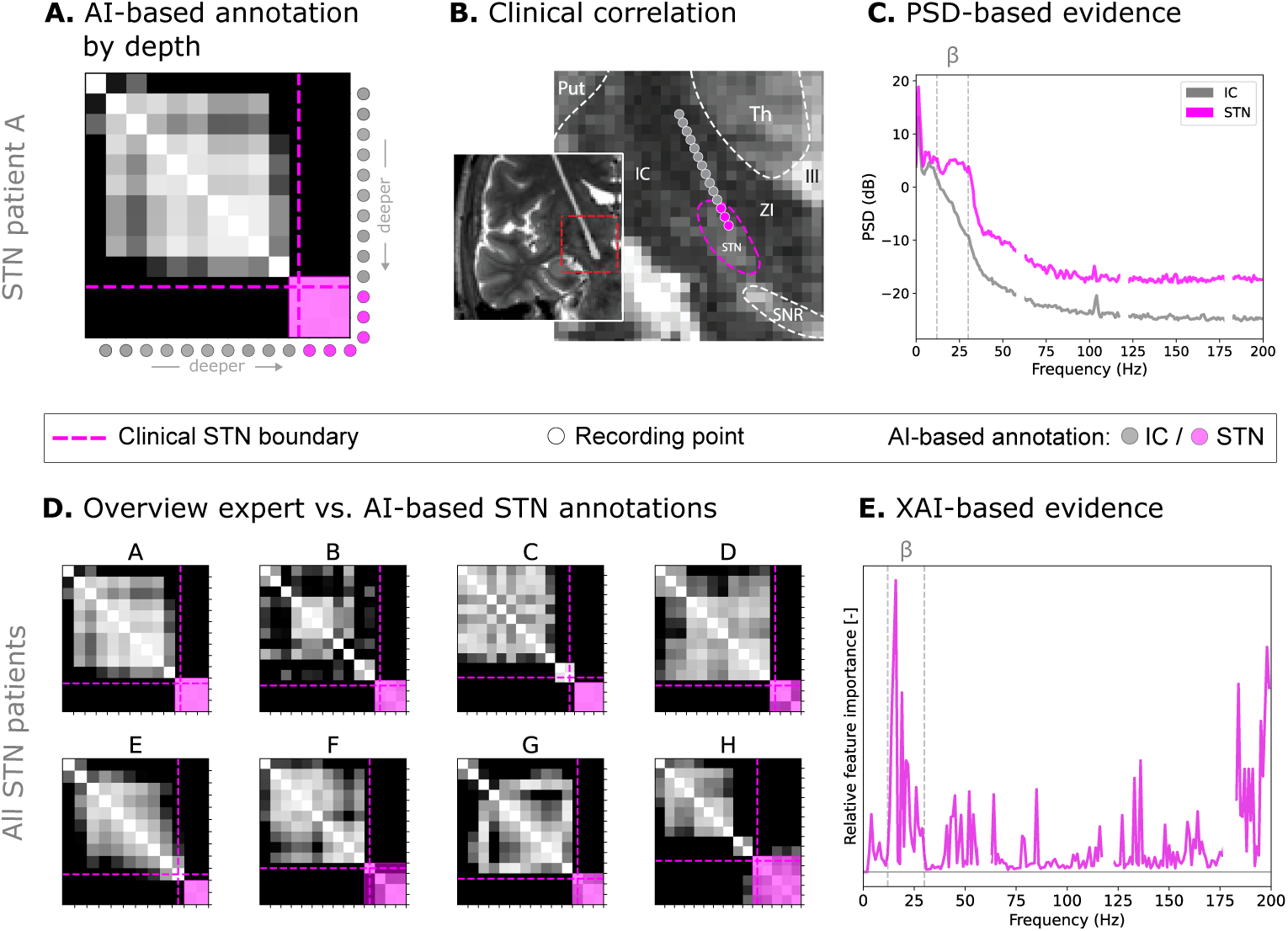
Algorithm validation on subthalamic nucleus (STN) detection. Detailed results for STN patient A in top row and overview of all STN patients in bottom row. **(A)** shows the imaging-based expert annotation of STN relative to the measurement locations of the DBS electrode. Locations are color-coded for being outside (grey) or inside (pink) the STN; neighboring regions are annotated for orientation. **(B)** The AI algorithm’s depth predictions result in clean clusters. The bottom cluster (pink) aligns exactly with the imaging-based expert annotation for the STN. **(C)** shows the PSD of raw signals averaged for measurements inside the STN (pink) and outside (grey). Signals from inside the STN show a distinct increase in beta-oscillations compared to the higher sites. **(D)** shows that the model was able to correctly identify the STN boundaries across all tested STN patients. Finally, **(E)** shows which spectral features the model used to distinguish between white matter and the STN (averaged across all patients). The analysis reveals that the model has mainly focused on the beta-range alongside some evidence for broadband oscillations. This validates that the model has learned a reasonable strategy to distinguish between STN and non-STN sites. Th = thalamus; IC = internal capsule; ZI = zona incerta; SNR = substantia nigra reticulata; Put = putamen; III = third ventricle.

### Discovering boundaries in the motor thalamus

We applied ThalaMAP to a cohort of 17 patients receiving Vim DBS for drug-resistant tremor. While the transition from the IC into the Vop is generally discernable on MRI and MER, the subsequent internal motor thalamus boundary between the Vop and Vim cannot be reliably visualized and lacks robust neurophysiological biomarkers (Fig. 1A-B).

For thalamus patient A, ThalaMAP reveals 3 distinct clusters along the trajectory (Fig. 3A). The first transition between the yellow and teal cluster perfectly aligns with the clinical annotation of the motor thalamus entry from white matter (green dashed line). The second transition reveals an internal thalamic boundary between the Vim and Vop, which is not apparent on MRI (Fig. 3B). We calculated the PSDs for each cluster and observed several differences in low frequency oscillations and broadband power (Fig. 3C).

**Fig. 3.**
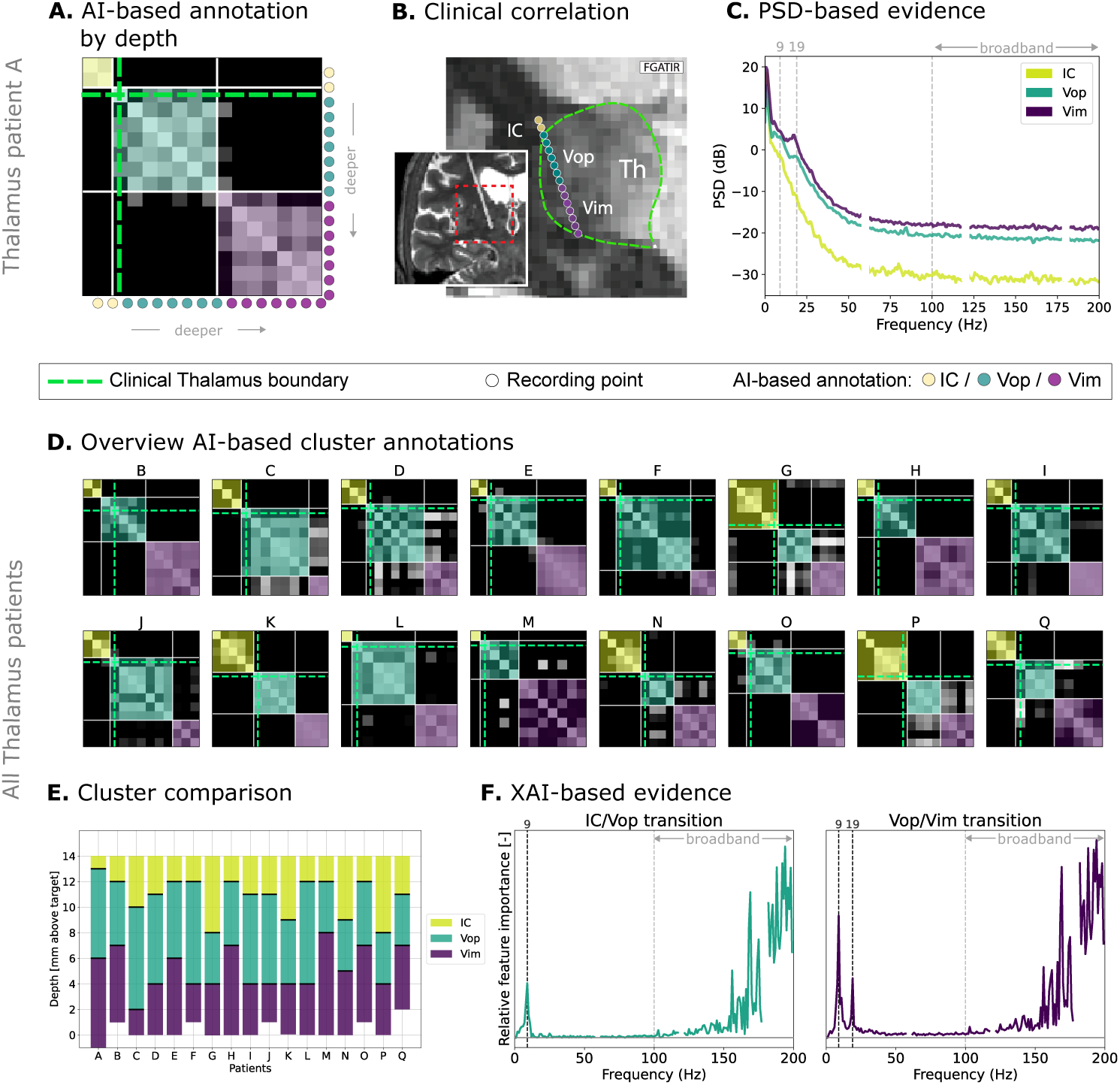
Overview of thalamus segmentation results. **(A)** This is an example trajectory from patient A. The magnification of the outlined windows displays the respective recording locations as well as the clinical annotation of the thalamus boundary. **(B)** Distance matrix of the respective depth predictions of patient A. Clusters manually (color-coded) annotated by domain experts based on model outcome. **(C)** Comparison of average power spectral densities for each resulting cluster for patient A. **(D)** Distance matrices and resulting clusters for all other patients (B to Q). **(E)** Cluster size and model-determined transition sites for all patients **(F)** Visualization of what frequencies were relevant for the model decision between the respective clusters. Vim = ventral intermediate nucleus of the thalamus; Vop = ventral oralis posterior; Th = thalamus; IC = internal capsule.

Remarkably, we find these three clusters consistently across all remaining 16 subjects with nearly perfect alignment of the annotated entry to the thalamus (Fig. 3D). The Vop cluster (green) is found on average at 10.9 *±* 1.5 mm above the target and spans over 5.8 *±*1.5 mm along the trajectory (compare Fig. 3E). The Vim cluster (purple) is entered at an average depth of 5.1 *±*1.6 mm above the target and traversed for 4.8 *±*1.6 mm, consistent with diffusion tractography estimates [18] (note, that the macroelectrode ring usually doesn’t reach the Vim ventral border). Cohort-level XAI relevance scores [15, 17] highlighted that the model detected the thalamus entry based on 9Hz oscillations and higher frequencies (*>*100), reflecting broadband power (Fig. 3F). For the internal thalamus transition (Vop/Vim) the 9Hz peak increases in relative importance and is accompanied by a secondary 19Hz centered peak. However, again broadband phenomena play a dominant role for the model’s ability to distinguish between the two regions. These features likely reflect the differing basal ganglia and cerebellar inputs to these subregions, revealing circuit-specific dynamics and local synaptic and spiking activity, providing objective intraoperative markers of thalamic organization. Detailed patient-specific PSD’s and evidence plots are presented in Appendix B. Notably, our framework performed significantly better than direct clustering of PSDs Appendix C.

### Functional validation using monopolar review

To evaluate whether physiologically defined clusters correspond to clinically meaningful differences in stimulation response, we analyzed postoperative monopolar review thresholds across contacts assigned to putative Vop and Vim clusters (Fig. 4A-E). Monopolar review represents the first systematic clinical assessment of stimulation effects after DBS implantation. It is a standardized clinical programming procedure in which vertical contact levels are tested individually while stimulation amplitude is gradually increased to evaluate therapeutic benefit (e.g. tremor reduction) and stimulation-induced side effects. this approach provides a systematic framework to characterize how different anatomical locations respond to stimulation in real patients.

**Fig. 4.**
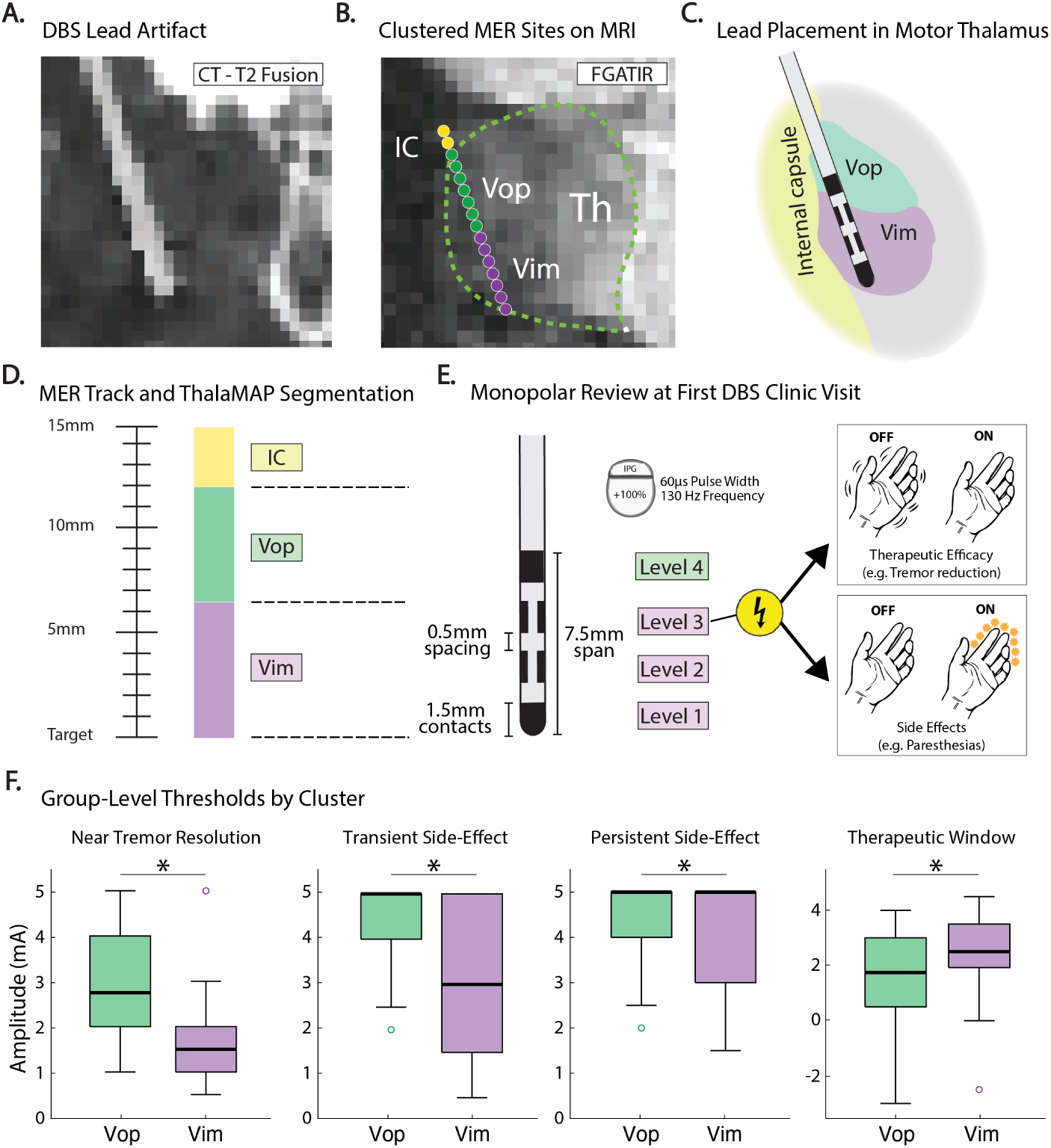
Physiologic cluster identity predicts stimulation-response properties during monopolar review. **(A)** Example of deep brain stimulation (DBS) lead artifact visualized on fused postoperative CT and preoperative MRI. **(B)** Clustered intraoperative microelectrode recording (MER) sites projected onto FGATIR MRI demonstrate physiologic segmentation along the trajectory, identifying internal capsule (IC), ventral oralis posterior (Vop), and ventral intermediate (Vim) regions. **(C)** Schematic representation of DBS lead placement within the motor thalamus, highlighting the spatial relationships between Vop, Vim, and adjacent structures. **(D)** MER trajectory annotated with ThalaMAP-derived segmentation showing physiologic boundary transitions along the electrode path. **(E)** Monopolar review workflow during the initial DBS clinic visit, in which individual contacts are tested to determine therapeutic efficacy thresholds and side-effect onset. **(F)** Group-level stimulation thresholds by physiologic cluster. Box-and-whisker plots display contact-level distributions for near-resolution threshold, transient side-effect onset, persistent side-effect onset, and calculated therapeutic window. Vim-associated contacts demonstrated lower efficacy thresholds and differences in side-effect thresholds compared with Vop, indicating cluster-dependent stimulation-response profiles. Boxes indicate interquartile range with median lines; whiskers extend to 1.5× the interquartile range, with points beyond this range shown as outliers. *indicates *p <* 0.05 for fixed effect of cluster ID.

If the AI-derived clusters reflect true functional organization, stimulation thresholds would be expected to differ between regions. Statistical comparisons were performed using linear mixed-effects models with cluster identity as a fixed effect and patient as a random effect to account for repeated measurements across contact levels. Contacts classified within the Vim cluster demonstrated significantly lower amplitudes required to achieve near-resolution of tremor compared with Vop-associated contacts (*F* = 26.60*, p* = 2.5610*^−^*6; *β* = *−*1.38 *±* 0.27*mA*), consistent with Vim representing the primary therapeutic target for tremor control (Fig. 4F). Transient side-effect onset also differed significantly between clusters (*F* = 17.07*, p* = 0.00010; *β* = *−*1.47 *±* 0.36*mA*), with lower thresholds observed in Vim-associated contacts, consistent with proximity to adjacent sensory thalamic regions where stimulation-induced side effects commonly arise [19]. Persistent side-effect thresholds showed a similar, but smaller cluster effect (*F* = 4.51*, p* = 0.037; *β* = *−*1.01 *±* 0.48*mA*). When therapeutic efficacy and side-effect thresholds were considered jointly, calculated therapeutic window (near resolution threshold - persistent side effect threshold) differed significantly between clusters (*F* = 5.95*, p* = 0.017; *β* = 0.87*±*0.36*mA*), with Vim-associated contacts demonstrating a broader therapeutic window relative to Vop.

Together, these findings indicate that AI-defined physiologic clusters derived from intraoperative LFP recordings correspond to distinct stimulation-response profiles observed during clinical programming, linking electrophysiologic boundary detection to functional neuromodulation outcomes.

## Discussion

As DBS and other neurosurgical therapies continue to expand for the treatment of neurological and psychiatric conditions, the ability to quantitatively define and target specific functional subregions becomes increasingly critical. Our findings demonstrate that integrating AI-driven analysis with traditional neurosurgical techniques can support more accurate, individualized targeting as well as tailored stimulation programming to improve patient outcomes. Supported by analysis of postoperative stimulation responses, our findings validate that AI-defined physiologic clusters exhibit distinct functional stimulation profiles, supporting their relevance for both optimal lead placement and subsequent programming decisions. This lays the foundation for developing intraoperative tools that provide real-time visualization of functional boundaries, aiding surgical decision-making. Furthermore, the ability to detect unique spectral profiles not only within the thalamus but potentially across other brain regions opens new opportunities for biomarker discovery and personalized neuromodulation strategies. Overall, this research advances both our understanding of thalamic physiological organization and the broader adoption of data-driven methods in neurosurgery, moving the field toward more precise and adaptive therapies for patients with complex brain disorders.

### Clinical Applications

After validation and offline testing, we deployed the ThalaMAP framework intraoperatively to evaluate its feasibility for real-time neurosurgical integration. Patient Q (Fig. 3D) underwent Vim DBS implantation for medication-refractory essential tremor. The model, trained on data from patients A–P, was applied directly to the macroelectrode LFP recordings acquired at 12 serial depths. Within seconds, ThalaMAP identified a transition from white matter of the internal capsule (IC) into the thalamus that precisely matched the boundary annotated intraoperatively by the surgical team based on single-unit firing patterns. A subsequent functional transition corresponding to the Vop/Vim border also clearly emerged, and the operating neurosurgeon confirmed the usefulness of this physiologic landmark in guiding final lead placement. Beyond intra-operative targeting, physiologic cluster identity also predicted stimulation-response properties during monopolar review at first DBS programming visit post implantation surgery. Contacts classified within Vim and Vop clusters demonstrated distinct efficacy and side-effect thresholds, consistent with established functional anatomy of the motor thalamus [9, 20].

These findings suggest that physiologic boundary detection may inform not only lead placement but also subsequent programming strategies, providing a bridge between intraoperative physiology and longitudinal clinical management. These results demonstrate the feasibility of integrating our framework into neurosurgical workflows and its potential value as a complementary, real-time resource for intraoperative decision-making. By converting serial LFPs into visualizable maps of functional boundaries, the system provides an objective counterpart to conventional physiological mapping and could help assist expert neurologist and neurophysiologist interpretation of brain signals. More broadly, this capability to detect otherwise invisible transitions underscores the potential of data-driven physiological mapping to enhance surgical precision, streamline workflows, and ultimately improve clinical outcomes in DBS and other functional neurosurgical procedures.

### XAI-derived Insights

Beyond boundary detection, our XAI analysis uncovered distinct spectral peaks around 9 Hz and 19 Hz along with broadband activity between Vim/Vop clusters. The lower-frequency peaks fall within the alpha and low–beta range, classical bands implicated in sensorimotor networks [21–23]. Their localization to the thalamic clusters suggests that rhythmic dynamics differentiate cerebellar (Vim) versus basal ganglia (Vop) input territories. The broadband components likely reflect population spiking or synaptic activity, with an increase in the Vim. Clinically, these physiologic markers may serve as objective intraoperative landmarks to guide electrode placement. In a wider context, the ability to extract physiologically interpretable features from intraoperative data establishes a framework for integrating functional circuit information into precision neurosurgery and the development of adaptive neuromodulation strategies.

### Limitations

Validating functional boundaries within the thalamus remains inherently challenging due to the absence of a definitive ground truth for Vim targeting. Unlike structures like the STN, where MRI visibility and well-defined physiological markers support clear boundary identification, the internal architecture of the motor thalamus lacks distinct imaging features, making direct comparisons between our model’s outputs and standard clinical practice difficult. Future efforts could compare model-derived boundaries to patient-specific anatomical reconstructions using co-registered brain atlases[24] or other advanced imaging methods [25], however atlas-based approaches have their own limitations and are not ground truth [6]. Encouragingly, application of the model to the STN accurately recapitulated known boundaries and demonstrated that the method can reliably detect physiologically meaningful transitions. During functional stimulation testing, monopolar review provides only a course vertical sampling (1.5mm per level), and individual stimulation levels may span adjacent physiologic clusters in some cases.

The annotation of clusters itself was mostly ubiquitous since the majority of patients exhibited clear, sharp transitions between clusters. In a few cases, however, boundaries were less distinct with potential minor shifts in cluster separation. This may be the case if boundaries are a gradient of overlapping inputs from different circuits resulting in a ‘blurry’ boundary depending on the spatial sampling [26]. We also note that, as generally the case for data-driven approaches, data quality is key for high quality model outcomes. In future iterations, boundary annotation could be further standardized by applying clustering methods to the distances of model output distributions [27]. Future work could evaluate whether patients with clearer physiologic boundary separations exhibit more favorable stimulation-response profiles or broader therapeutic windows during clinical programming.

### Future Directions

Building on these findings, future work will extend this machine learning framework to additional brain regions beyond the thalamus and STN, including targets such as the internal globus pallidus and anterior limb of the internal capsule, where precise functional boundaries remain difficult to define [28]. Incorporating additional neural signals such as spiking activity, evoked potentials, and high-frequency oscillations may further improve boundary detection and expand use-cases. Beyond anatomical mapping, these methods could identify state-dependent physiological patterns, offering applications in emerging closed-loop DBS systems [29–32].

While our model reliably identified clusters corresponding to the Vop and Vim subregions and demonstrated cluster-dependent differences in stimulation-response properties, future work should evaluate whether AI-derived boundary information can prospectively inform targeting strategies and guide individualized programming configurations. The current use of serial MER introduces limitations related to temporal stability, as patient state and LFP dynamics may fluctuate over time. Incorporating high-density or array-based microelectrodes capable of simultaneous multi-site recordings along the lead trajectory could mitigate these effects, providing more stable, spatially precise, and temporally rich data. Such parallel recordings would also enable advanced temporal analyses, including phase-based coherence measures [33]. Finally, expanding our dataset to include a larger and more diverse patient population will be essential to assess inter-subject variability and enhance model generalizability across broader clinical contexts.

## Materials and Methods

### Ethics statement

The study was conducted according to the Institutional Review Board of the Mayo Clinic (IRB 19-009878), which also authorizes sharing of de-identified data. Each patient provided informed written consent as approved by the IRB. Patient MRI sequences were anonymized by only including slices in-plane to the DBS leads for visualization.

### Subjects

DBS leads were placed into the ventral Vim nucleus of the thalamus in 17 patients (11 males, 6 females) for treatment of medication-refractory essential tremor. For the validation cohort, leads were placed in the STN of 8 patients (6 males, 2 females) for treatment of medication-refractory Parkinson’s Disease. Both cohorts participated in a research protocol involving serial microelectrode recordings along the planned trajectory of the DBS lead.

### Recordings

Serial thalamic recordings were obtained at 1 mm apart evenly spaced recording sites as a hybrid microelectrode/macroelectrode (AlphaOmega Sonus STR-009080-00) was advanced towards a target at the inferior border of the thalamus (Fig. 1A). Voltage data were recorded from the macroelectrode ring (+3mm from the tip) and referenced to the shaft of the electrode cannula with sampling rate of 44 kHz. At each depth, recording commenced 2 seconds after electrode movement stopped and continued for approximately 30 seconds. No somatosensory or visual evoked potential or micro/macro-stimultation testing were included in the analyzed data.

### Localization of recording sites

As the serial recordings followed the same trajectory as the permanent DBS lead, each depth could be localized relative to the final lead position. Localization entailed:

1) Co-registration of the postoperative CT scan to the preoperative MRI sequences using the normalized mutual information approach in SPM12 [34]) manual identification of lead’s contact artifacts in the postoperative CT (GOV), and 3) adjustment for the known distance of each recording depth from the identified positions the DBS lead contacts. To optimize visualization, MRI sequences were resliced in plane with the lead using publicly available custom Matlab code [14]. Recording depths were then plotted on T1-weighted, T2-weighted, and Fluid-attenuated Gradient echo with Adiabatic T1 Inversion Recovery (FGATIR) MRI sequences. MRI borders of the thalamus and surrounding structures were visually identified and outlined in the figures. See Appendix D for expanded schematic on MER recording, lead localization, and visual anatomical segmentation.

### Signal processing

The signal processing pipeline from the raw recording to the model input consists of several consecutive filtering, transformation and scaling steps. First, we removed measurements with less than 20 seconds of recording time (7 in total) to ensure homogeneity of the recording setting and a balanced data set. The recordings at each depth level of the 17 individuals were divided into 1-second windows (with 25% overlap) and transformed to the frequency domain by discrete Fourier Transform [35] (Fig. 1C). We restrict the resulting feature space to frequencies from two up to 200 Hz, where we can capture physiologically meaningful narrow-band oscillations and broadband features [36–38]. We mask 60 *±*2 Hz harmonics to remove potential line noise contamination. As a result we obtain approximately 6000 samples, each with an annotated recording depth, which serve as a self-supervised regression target. For our study, we divide our datasets into training and test sets in a leave-one-patient-out fashion (16 training, 1 test). Thus, the model is trained on data from all other patients and applied to the respective left out patient to assess model generalization across individuals, with 20% of the training samples randomly selected and set aside as a validation set for earlystopping.

### AI model

We utilize a vanilla multilayer perceptron (MLP) with ReLU-activations and five fully connected layers (10 neurons each, *≈* 2500 learnable parameters in total) (Fig. 1C). The model is trained using the Adam algorithm [39] (initial learning rate of 0.01 scheduled to reduce if the training error does not reduce further, random weight initialization) and model inputs are min-max scaled by frequency bin to facilitate the optimization. In addition to the mean squared error loss, we apply L2-regularization (*λ_L_*_2_ = 0.01) and additionally introduce an L1-loss term (*λ_L_*_1_ = 1). The latter is applied to the weights of the first layer only and therefore represents an implicit feature selection which forces the model to focus on the most important frequencies when mapping a sample to its recording depth. This results in more sparse explanations when attributing the model decision to the respective input features with XAI attribution methods [16, 40], and therefore facilitates the identification of biomarkers (compare Appendix E for results without the L1-penalty term). In summary, we optimize the following training objective over the number of training samples *n*:

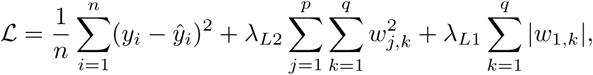

where *y_i_*and *ŷ_i_* represent the true and predicted recording depth of sample *i*, *p* the number of model layers and *q* the number of neurons per layer (*w_j,k_* being the weight *k* in layer *j*, respectively).

During training we employ early stopping if the validation error has not reached a new minimum within the previous 100 epochs. Training is usually terminated after approximately 1000 epochs corresponding to around 30 seconds on a standard laptop. Once the model has been trained, inferences can be made in real-time (sub-second).

### Model output post-processing

To effectively identify functional boundaries of each individual patient, we post-process the model’s depth predictions. First, we group them according to the true depth and obtain the conditional distribution *p*(*ŷ_model_|y_true_*). Then, we calculate first-order, non-parametric Wasserstein-distances [41] between these distributions for each pair of recording depths. In the given case this reduces to the computationally efficient calculation of the mean absolute difference between sorted model outputs per recording depth. Visualizing the resulting pairwise distances in a matrix plot enables the immediate identification of clusters which appear as block structures along the diagonal. The transitions between the blocks (sub-regions) were manually annotated by analyzing the distance matrices as well as the comparison of kernel density plots of the respective predictions (Fig. 1D-E1). Identified clusters can then be projected back onto microelectrode recording sites with respective colors (Fig. 1E2).

### Explainable AI for biomarker discovery

To better understand which spectral features are relevant for the model to distinguish between clusters, we use layer-wise relevance propagation (LRP) [15, 16], a popular method from the field of explainable AI (XAI). LRP propagates the output of a ML model back through its architecture and results in relevance attributions for each input feature (e.g. how relevant was feature *i* for the output of sample *j*). We use the standard LRP-0 rule and its implementation from the Zennit library [42]. We further aggregate these individual attributions across predicted depths and patients to gain insights into the overall learned model strategy [17, 43]. Concretely, we calculate the average attributions for each cluster across patients and normalize them to values between zero and one. Then, we subtract the averaged cluster attributions from each other to focus on the contrastive question of what frequency feature is important for the distinction between cluster A and cluster B. The resulting explanations can be utilized in two distinct ways. For regions where concrete frequency-based biomarkers are known, we can validate whether our model has learned to utilize the ‘expected’ information for its decision. For brain regions where clinical biomarkers are not as well understood, the explanations can be a starting point to guide us towards the potential discovery of new biomarkers [40, 44, 45].

### Clinical monopolar review and stimulation analysis

Postoperative clinical programming data were obtained from standardized monopolar review sessions performed during each patient’s first DBS follow-up visit several weeks following lead implantation. During monopolar review, individual vertical contact levels were tested sequentially using fixed stimulation parameters (60µs pulse-width and 130Hz frequency) while stimulation amplitude was gradually increased to assess therapeutic efficacy and stimulation-induced side effects. Each stimulation level corresponds to an axial depth along the DBS lead; for directional leads, multiple segmented contacts at the same depth (e.g., contacts 2–4 or 5–7) were treated as a single stimulation level for analysis. For each level, amplitude thresholds were recorded for near-resolution of tremor, transient side-effect onset, and persistent side-effect onset. A calculated therapeutic window was defined as the difference between efficacy threshold and persistent side-effect threshold.

Levels were assigned to physiologic clusters (Vop or Vim) based on their spatial correspondence to ThalaMAP-derived segmentation along the electrode trajectory. In cases where near-resolution of tremor or stimulation-induced side effects were not observed within the tested amplitude range, threshold values were assigned a maximum value of 5 mA. This reflects inherent clinical constraints during monopolar review, where amplitude escalation may be limited by tolerability or other clinical considerations. Statistical comparisons between clusters were performed using linear mixed-effects models with cluster identity as a fixed effect and patient as a random effect to account for within-subject correlations. Statistical significance was defined using a two-tailed alpha threshold of 0.05.

## Data Availability

A provisional patent application has been filed related to the methods described in
this work. Custom code developed for this study will be made available to academic
researchers for non-commercial use under a research license. The code will be hosted at https://github.com/sltzgs/ThalaMAP. Access and licensing inquiries should be directed to the corresponding authors.

## Data and code availability

A provisional patent application has been filed related to the methods described in this work. Custom code developed for this study will be made available to academic researchers for non-commercial use under a research license. The code will be hosted at https://github.com/sltzgs/ThalaMAP. Access and licensing inquiries should be directed to the corresponding authors.

## Acknowledgements

This work was supported by the National Institutes of Health (NIH) NINDS F31-NS135898 (MAJ), NINDS U01-NS128612 (MRB, KJM). Manuscript contents are solely the responsibility of the authors and do not necessarily represent the official views of the NIH.

MRB, KJM, and BTK were supported by the MN partnership grant for biotechnology and medical genomics (MNP2142). MRB and KJM were also supported by a Helene Houle Career Development Award and the Tianqiao & Chrissy Chen Institute.

KRM was supported in part by the German Ministry for Education and Research (BMBF) under Grants 01IS14013A-E, 01GQ1115, 01GQ0850, 01IS18025A, 031L0207D, 01IS18037A as well as Berlin Institute for the Foundations of Learning and Data (BIFOLD). Furthermore KRM was partly supported by the Institute of Information & Communications Technology Planning & Evaluation (IITP) grant funded by the Korea government (MSIT) (No. RS-2019-II190079, Artificial Intelligence Graduate School Program, Korea University) and grant funded by the Korea government(MSIT) (No. RS-2024-00457882, AI Research Hub Project).

SL was supported in part by the German Ministry for Education and Research (BMBF) under Grant 16IS24087A.

The funders had no role in study design, data collection and analysis, decision to publish, or preparation of the manuscript. ChatGPT (OpenAI) was used for assistance in text editing but not content generation.

## Appendix A: Detailed results per Subthalamic Nucleus patient

Here, we report the detailed ThalaMAP results for each of the STN patients (A to H) in Figures 5 and 6. They complement the aggregated results shown in Figure 2. We show for each patient (from left to right): the scatter plot for all samples against their true label, the respective kernel-density plots for each predicted depth, the distance matrix between the distributions with annotated IC transition and clusters, the respectively color-coded PSDs from each raw recording per depth, and the explanations aggregated over the samples of the respective patient.

**Fig. 5.**
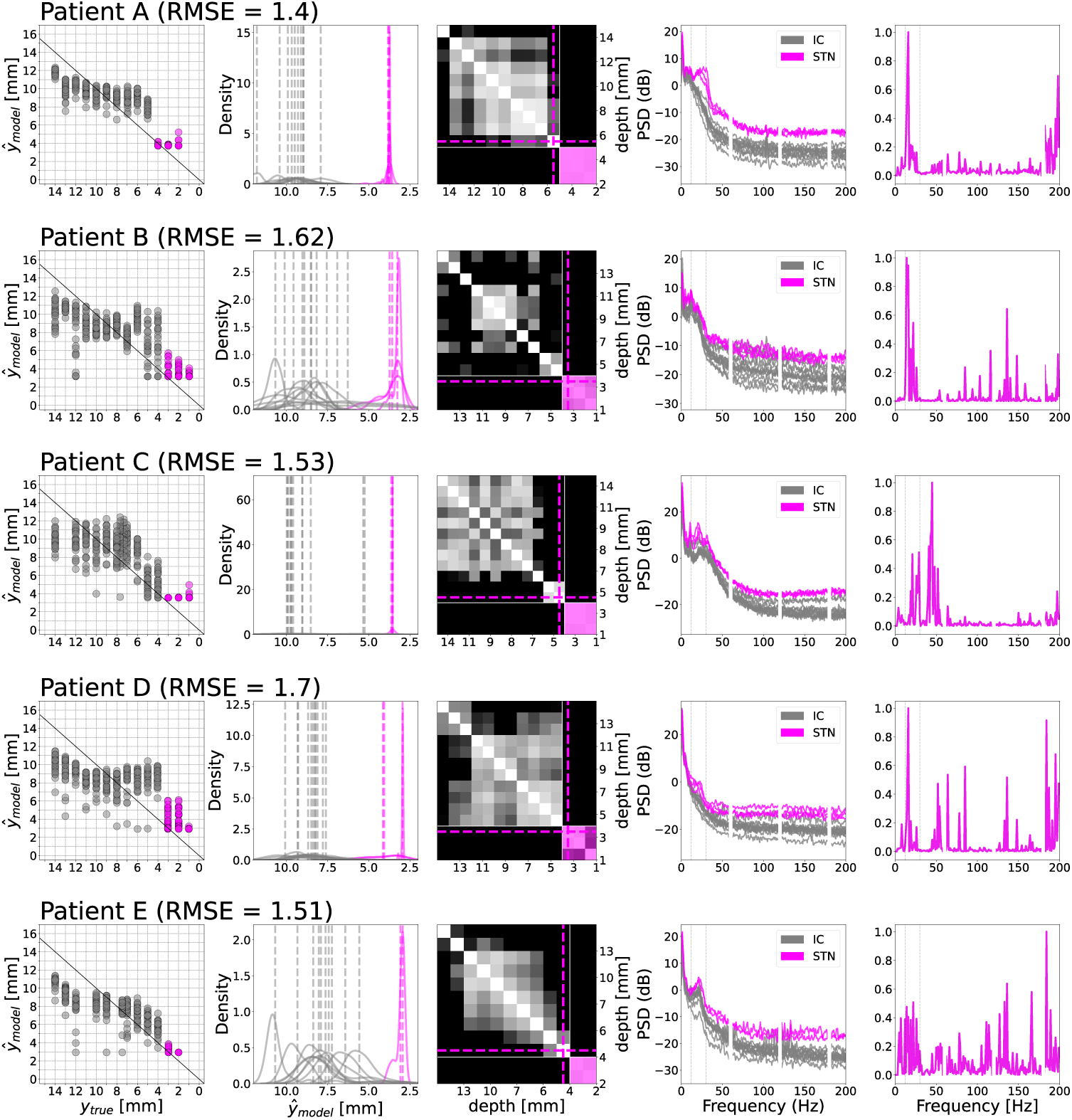
Detailed results for STN patients A to E complementing the results shown in Figure 2.

**Fig. 6.**
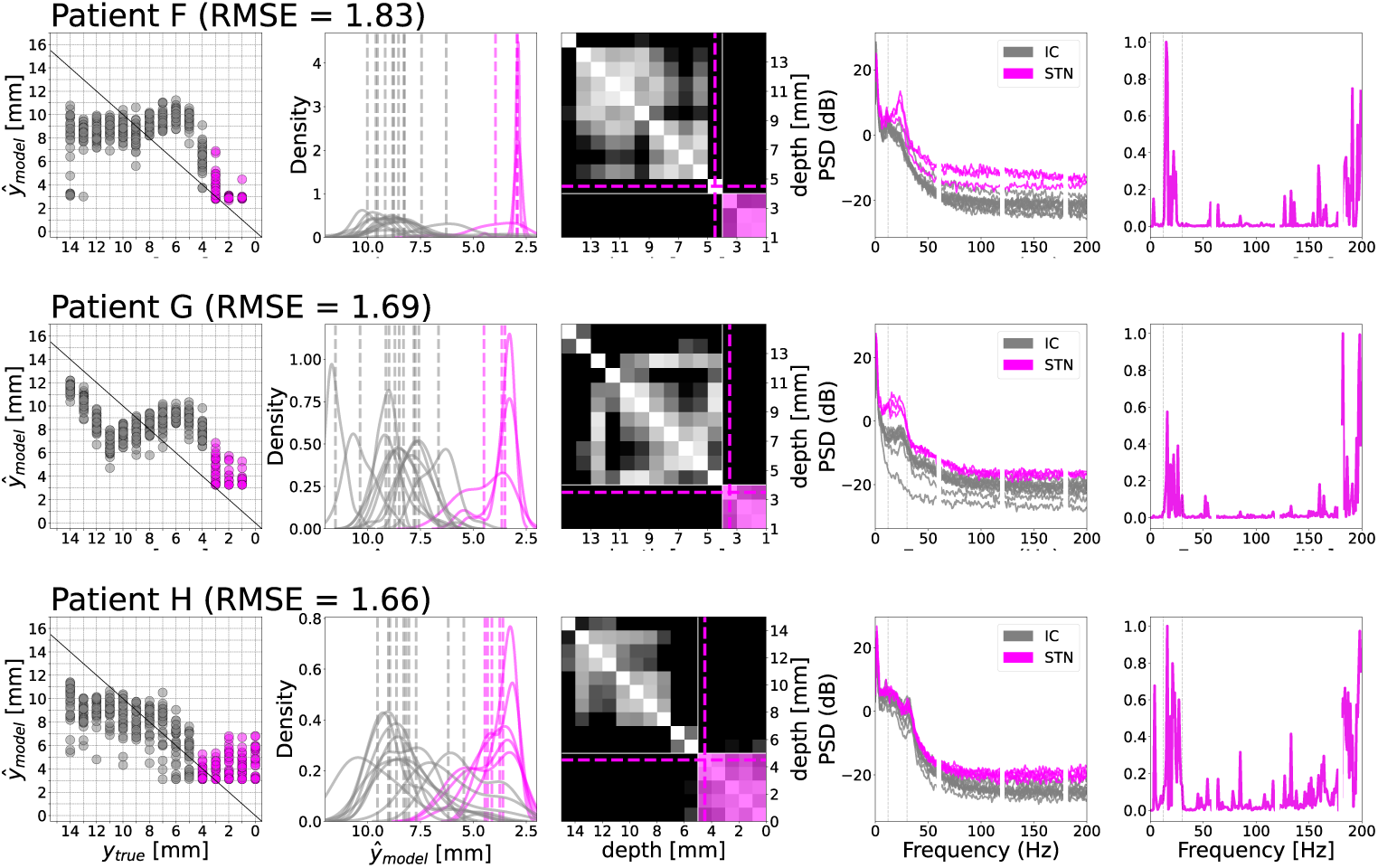
Detailed results for STN patients F to H complementing the results shown in Figure 2.

## Appendix B: Detailed results for all thalamus patients

Here, we report the detailed ThalaMAP results for each of the thalamus patients (A to Q). Figures 7 to 9 complement the aggregated results shown in Figure 3. We show for each patient (from left to right): the scatter plot for all samples against their true label, the respective kernel-density plots for each predicted depth, the distance matrix between the distributions with annotated IC transition and clusters, the respectively color-coded PSDs from each raw recording per depth, and the explanations aggregated over the samples of the respective patient.

**Fig. 7.**
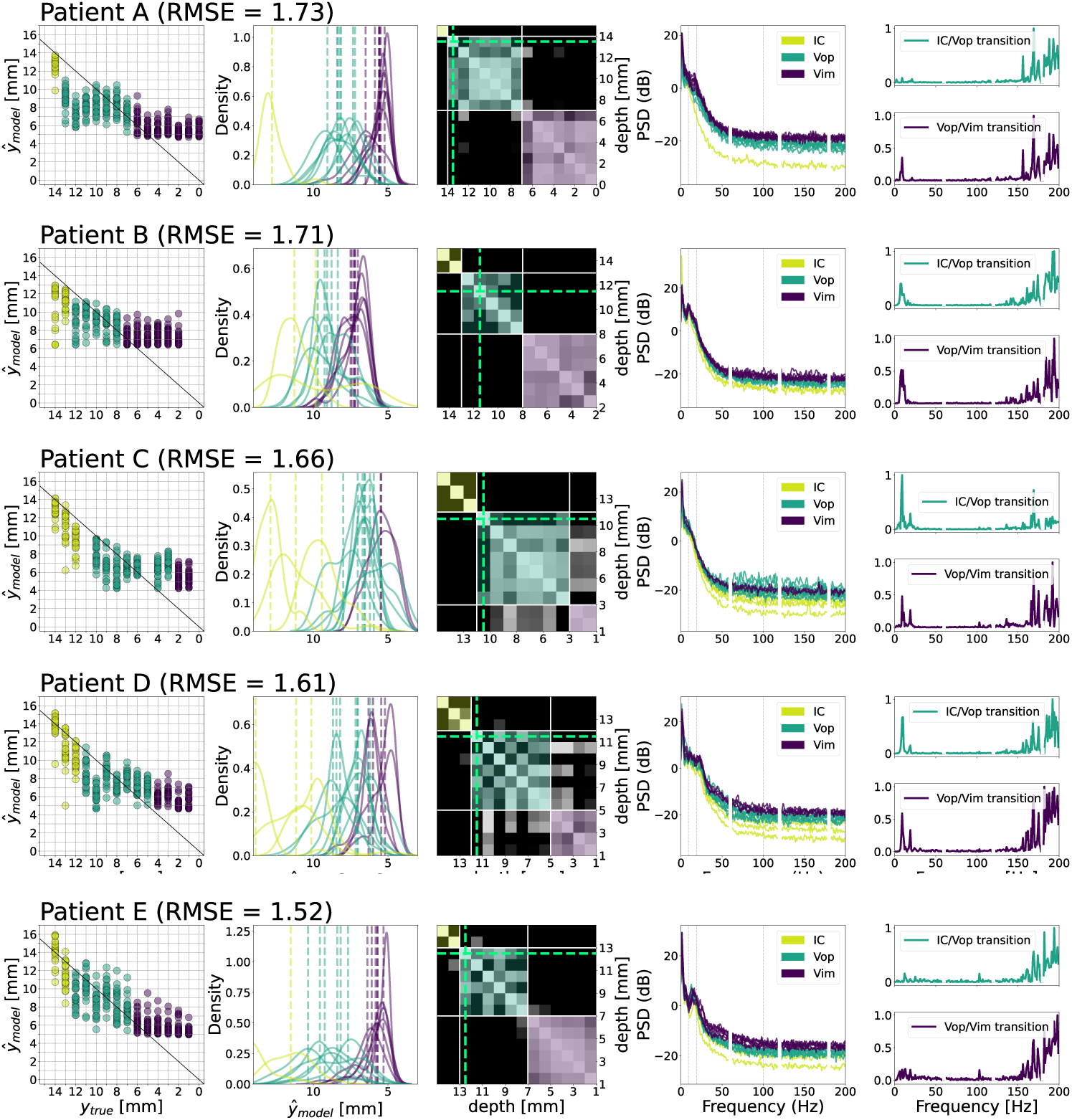
Detailed results for thalamus patients A to E complementing results shown in Figure 3.

**Fig. 8.**
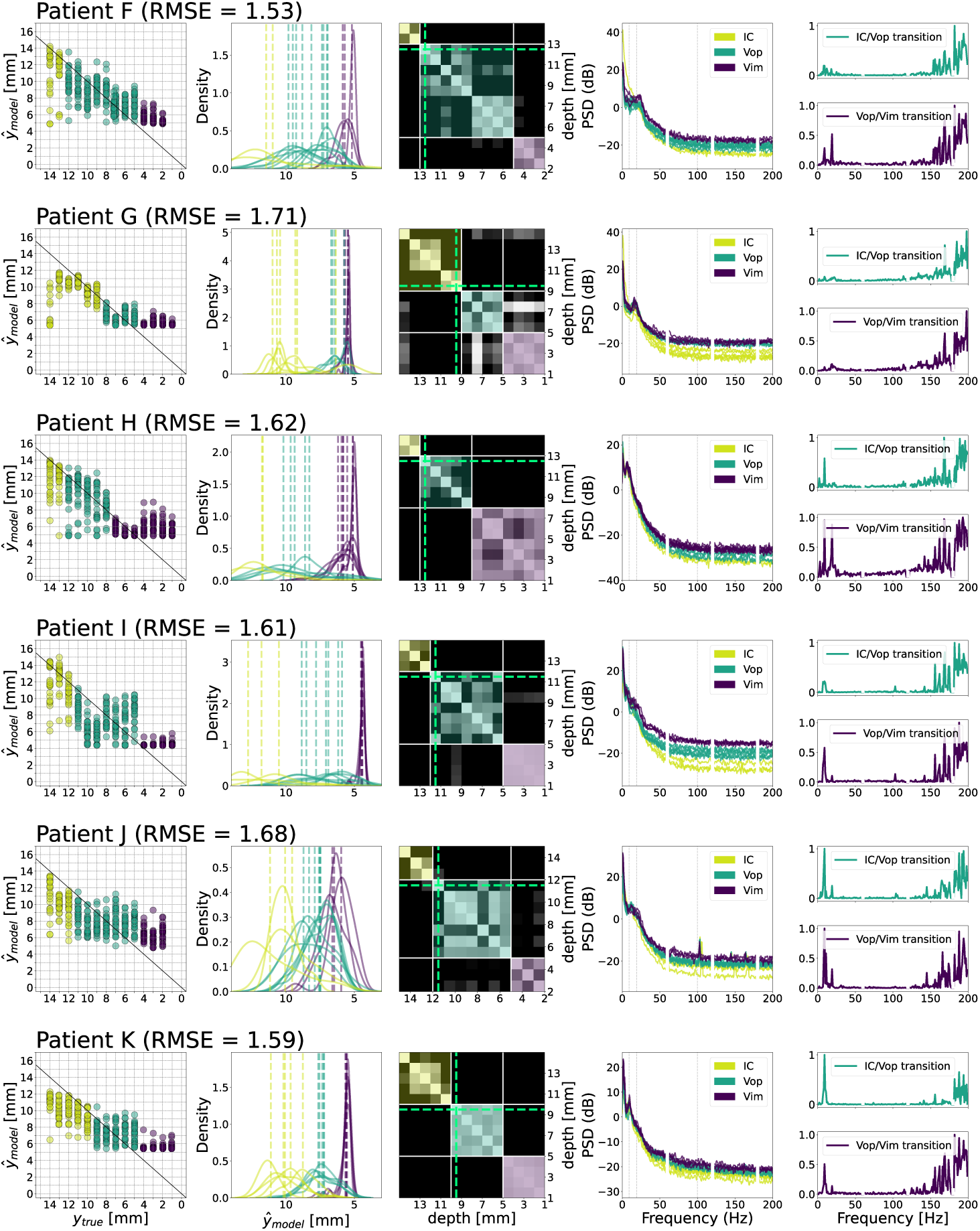
Detailed results for thalamus patients F to K complementing results shown in Figure 3.

**Fig. 9.**
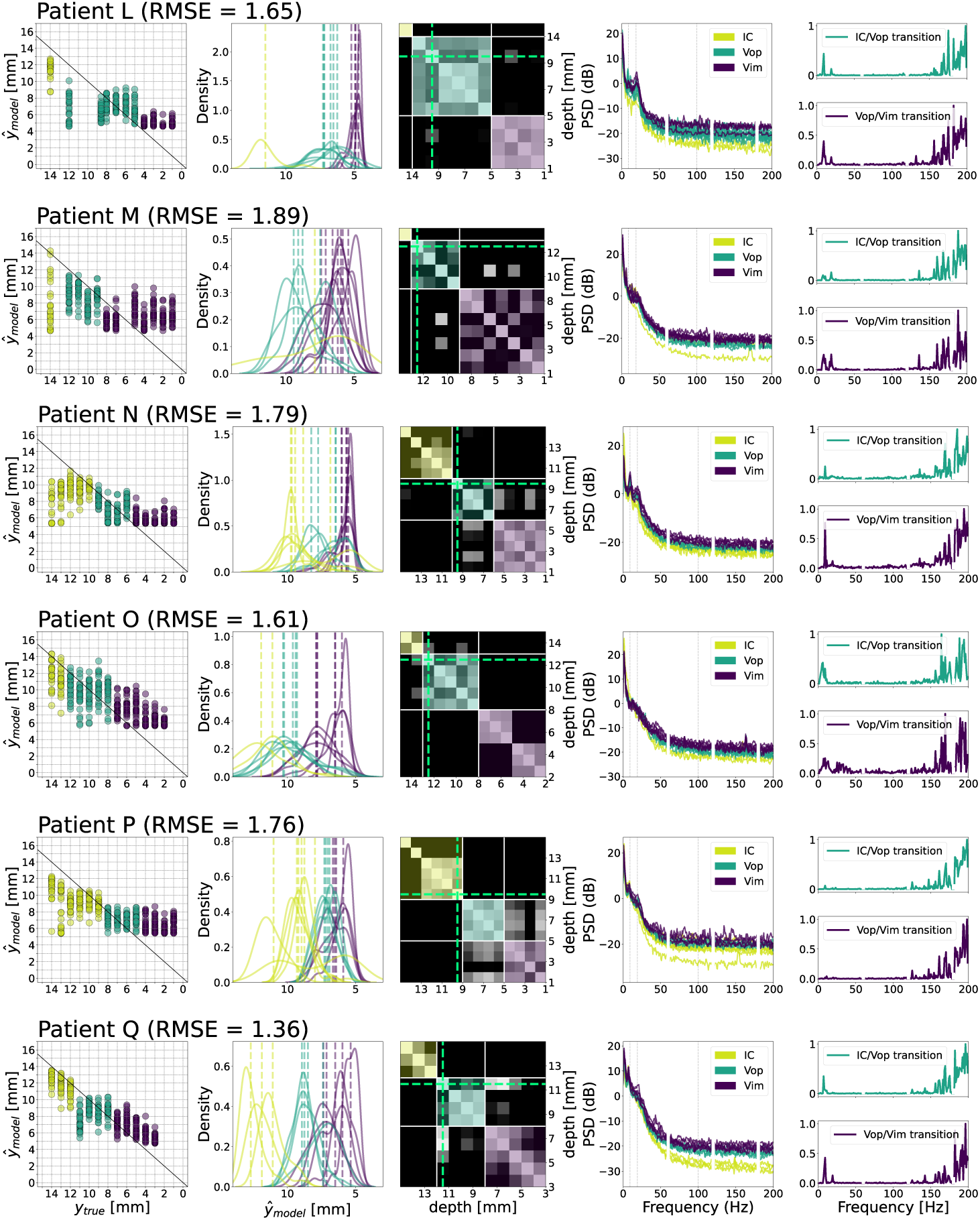
Detailed results for thalamus patients L to Q complementing results shown in Figure 3.

## Appendix C: Direct clustering of PSD representations

In this section, we show the comparison of our approach with direct clustering of the power spectral density of the respective recording depths for k-means (Fig. 10) and spectral (Fig. 11) clustering. We used the respective scikit-learn [46] implementations.

**Fig. 10.**
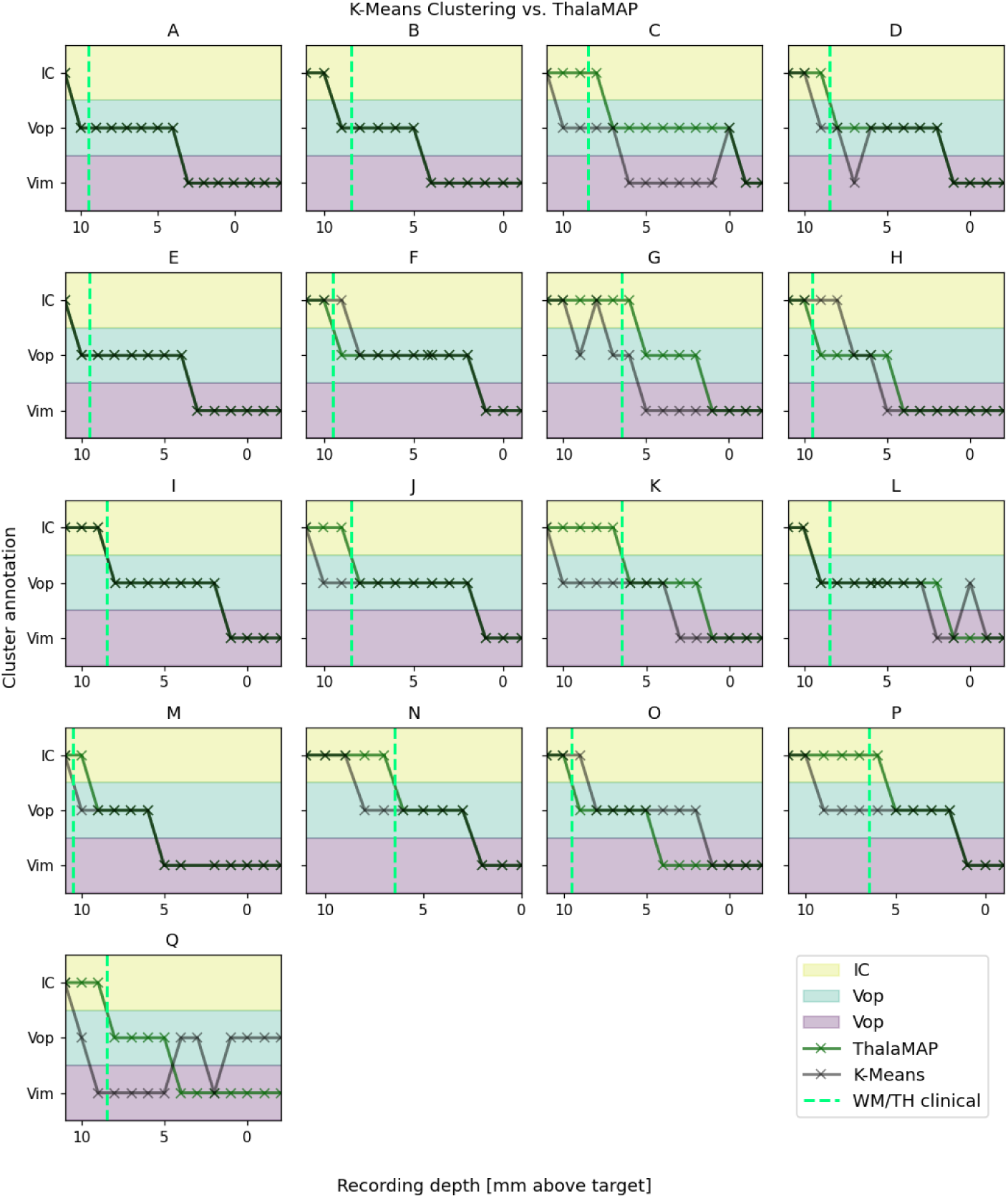
Comparison of K-means (black) and ThalaMAP (green). Y-axis shows cluster annotation, x-axis shows recording depth. The clinically determined IC/thalamus transition is shown as a dashed green line.

Figure 10 shows the cluster annotations of the k-means algorithm compared to ours (ThalaMAP) and the clinical white matter, thalamus (WM/TH) transition. We can see that the outcomes for patients A, B, E and I are perfectly aligned and therefore equivalent. For all others, however, we see clear deviations including many physiologically implausible annotations (reverse transitions). Moreover, the WM/TH transition is annotated poorly with errors up to 4 mm (patient K).

**Fig. 11.**
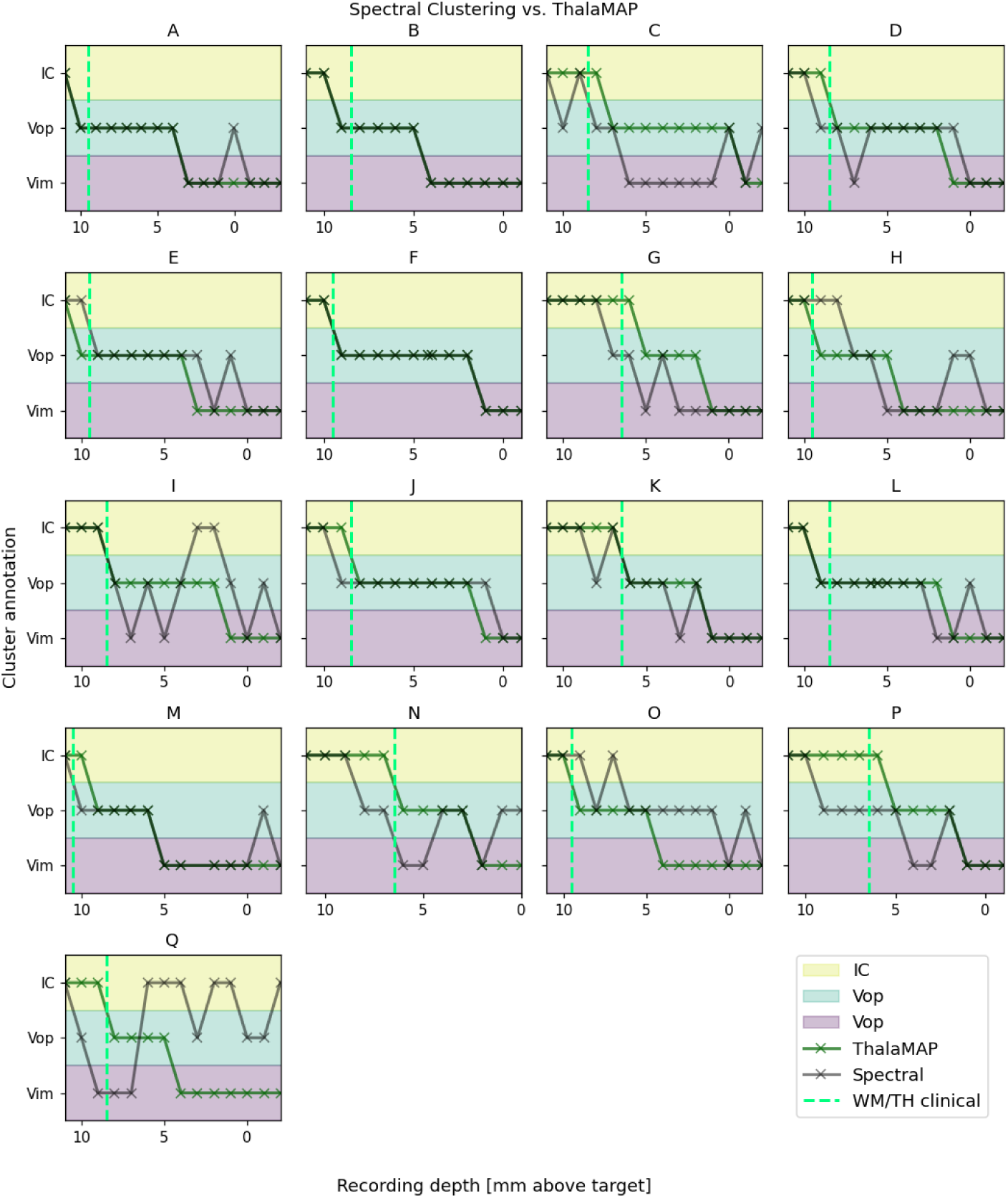
Comparison of Spectral Clustering (black) and ThalaMAP (green). Y-axis shows cluster annotation, x-axis shows recording depth. The clinically determined WM/thalamus transition is shown as a dashed green line.

Figure 11 shows the cluster annotations of the Spectral Clustering algorithm compared to ours (ThalaMAP) and the clinical White Matter, thalamus (WM/TH) transition. Here, we see perfect alignment in only two cases (B and F), while most others show physiologically implausible annotations (reverse transitions).

These findings demonstrate the superiority of the proposed approach over naive clustering baselines which can be attributed to incorporation of recording depth as a soft label. Moreover, the naive clustering approaches lack the ability to point to specific biomarkers.

## Appendix D: Intraoperative microelectrode recordings and localization

Here, we report an expanded schematic for microelectrode recording (MER) methods, lead localization, and visible anatomy segmentation. The microelectrode is passed along thalamic subregions and records local field potential data from the macroelectrode contact +3mm from the tip (Fig. 12A-B). DBS leads are localized on merged imaging and resliced in-plane with the lead to visualize the inferred recording track (Fig. 12C-D). Visible anatomical boundaries are hand-segmented on imaging (Fig. 12E).

**Fig. 12.**
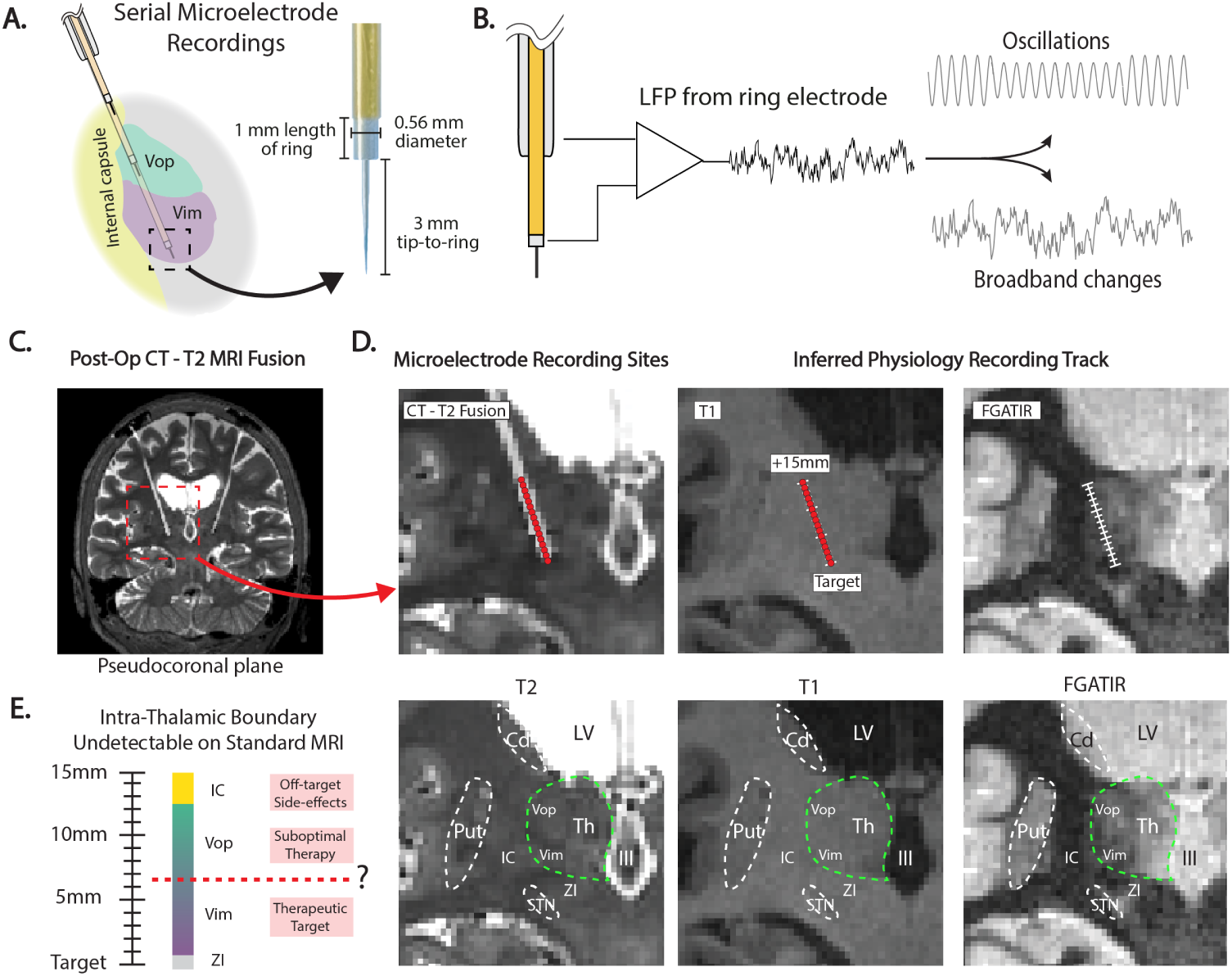
Intraoperative serial microelectrode recordings for deep brain stimulation (DBS) surgery targeting the ventral intermediate nucleus of the thalamus (Vim) for tremor. **(A)** Diagram depicting microelectrode being passed along thalamic subregions (coronal view) and microelectrode dimensions. **(B)** Local field potentials are recorded from the ring electrode and referenced to the cannula, revealing neural oscillation and broadband information. **(C)** DBS leads are localized on postoperative CT and T2 MRI fusion resliced in-plane with the leads. **(D)** DBS lead artifact is used to infer microelectrode recording locations. **(E)** Visible anatomical boundaries along the microelectrode recording track outlined on MRI. Notably, thalamic subregions are not visible on MRI. IC = internal capsule; VC = ventral caudal; Vim = ventral intermediate nucleus of the thalamus; Vop = ventral oralis posterior; Put = putamen; Cd = caudate; LV = lateral ventricle; III = third ventricle; Th = thalamus; ZI = zona incerta; STN = subthalamic nucleus.

## Appendix E: XAI explanation without L1-loss

Here, we report the ThalaMAP results for models trained without the L1-regularization term. While the overall cluster annotations remain the same (compare Fig. 13 and Fig. 3D), the XAI attributions are much less sparse and, therefore, much more difficult to interpret (compare Fig. 14).

**Fig. 13.**
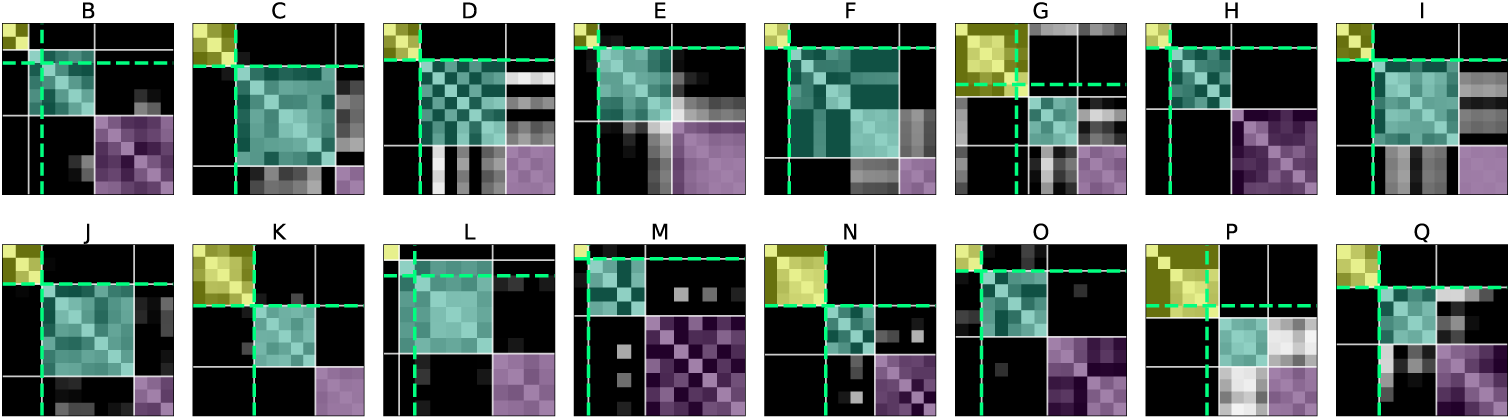
ThalaMAP results for models without the L1-regularization term. Overall cluster annotations remain the same as the results reported in section 7

**Fig. 14.**
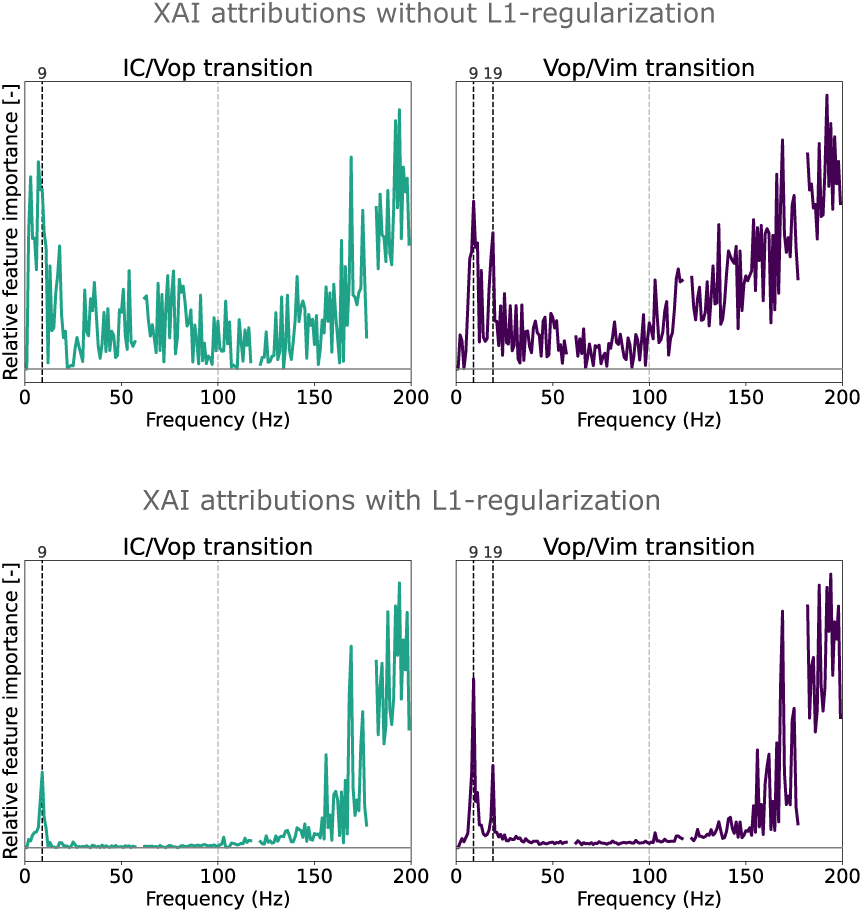
XAI attributions for transitions between regions when models are trained without the L1-regularization term (top) and with the L1-regularization term (bottom).

## Notes

### Competing Interest Statement

The authors have declared no competing interest.

## References

[1] Budman, E., Deeb, W., Martinez-Ramirez, D., Pilitsis, J.G., Peng-Chen, Z., Okun, M.S., Ramirez-Zamora, A.: Potential indications for deep brain stimulation in neurological disorders: an evolving field. European Journal of Neurology 25(4), 434–445 (2018)

[2] Lozano, A.M., Lipsman, N., Bergman, H., Brown, P., Chabardes, S., Chang, J.W., Matthews, K., McIntyre, C.C., Schlaepfer, T.E., Schulder, M., Temel, Y., Volkmann, J., Krauss, J.K.: Deep brain stimulation: current and future clinical applications. Nature Reviews Neurology 15(3), 148–160 (2019)

[3] Montgomery, E.B., Gale, J.T.: Mechanisms of action of deep brain stimulation (dbs). Neuroscience Biobehavioral Reviews 32(3), 388–407 (2008)

[4] Lehman, V.T., Lee, K.H., Klassen, B.T., Blezek, D.J., Goyal, A., Shah, B.R., Gorny, K.R., Huston, J., Kaufmann, T.J.: Mri and tractography techniques to localize the ventral intermediate nucleus and dentatorubrothalamic tract for deep brain stimulation and mr-guided focused ultrasound: a narrative review and update. Neurosurgical focus 49(1), 8 (2020)

[5] Su, J.H., Choi, E.Y., Tourdias, T., Saranathan, M., Halpern, C.H., Henderson, J.M., Pauly, K.B., Ghanouni, P., Rutt, B.K.: Improved vim targeting for focused ultrasound ablation treatment of essential tremor: A probabilistic and patient-specific approach. Human Brain Mapping 41(17), 4769–4788 (2020)

[6] Najdenovska, E., Tuleasca, C., Jorge, J., Maeder, P., Marques, J.P., Roine, T., Gallichan, D., Thiran, J.-P., Levivier, M., Bach Cuadra, M.: Comparison of mri-based automated segmentation methods and functional neurosurgery targeting with direct visualization of the ventro-intermediate thalamic nucleus at 7t. Scientific reports 9(1), 1119 (2019)

[7] Shanker, V.: Essential tremor: diagnosis and management. bmj 366 (2019)

[8] Ferreira, F., Akram, H., Ashburner, J., Zrinzo, L., Zhang, H., Lambert, C.: Ventralis intermedius nucleus anatomical variability assessment by mri structural connectivity. Neuroimage 238, 118231 (2021)

[9] Bosch-Bouju, C., Hyland, B.I., Parr-Brownlie, L.C.: Motor thalamus integration of cortical, cerebellar and basal ganglia information: implications for normal and parkinsonian conditions. Frontiers in computational neuroscience 7, 163 (2013)

[10] Koelman, L.A., Lowery, M.M.: Beta-band resonance and intrinsic oscillations in a biophysically detailed model of the subthalamic nucleus-globus pallidus network. Frontiers in Computational Neuroscience 13, 77 (2019)

[11] Weinberger, M., Mahant, N., Hutchison, W.D., Lozano, A.M., Moro, E., Hodaie, M., Lang, A.E., Dostrovsky, J.O.: Beta oscillatory activity in the subthalamic nucleus and its relation to dopaminergic response in parkinson’s disease. Journal of neurophysiology 96(6), 3248–3256 (2006)

[12] Bengio, Y., Courville, A., Vincent, P.: Representation learning: A review and new perspectives. IEEE transactions on pattern analysis and machine intelligence 35(8), 1798–1828 (2013)

[13] Doersch, C., Gupta, A., Efros, A.A.: Unsupervised visual representation learning by context prediction. In: Proceedings of the IEEE International Conference on Computer Vision, pp. 1422–1430 (2015)

[14] Richner, T.J., Klassen, B.T., Miller, K.J.: An in-plane, mirror-symmetric visualization tool for deep brain stimulation electrodes. In: 2020 42nd Annual International Conference of the IEEE Engineering in Medicine & Biology Society (EMBC), pp. 1112–1115 (2020). IEEE

[15] Bach, S., Binder, A., Montavon, G., Klauschen, F., Müller, K.-R., Samek, W.: On pixel-wise explanations for non-linear classifier decisions by layer-wise relevance propagation. PLoS ONE 10(7), 0130140 (2015) 10.1371/journal.pone.0130140

[16] Samek, W., Montavon, G., Lapuschkin, S., Anders, C.J., Müller, K.-R.: Explaining deep neural networks and beyond: A review of methods and applications. Proceedings of the IEEE 109(3), 247–278 (2021)

[17] Letzgus, S., Wagner, P., Lederer, J., Samek, W., Müller, K.-R., Montavon, G.: Toward explainable artificial intelligence for regression models: A methodological perspective. IEEE Signal Processing Magazine 39(4), 40–58 (2022)

[18] Kincses, Z.T., Szabó, N., Valálik, I., Kopniczky, Z., Dzsi, L., Klivnyi, P., Jenkinson, M., Király, A., Babos, M., Vörös, E., et al.: Target identification for stereotactic thalamotomy using diffusion tractography. PloS one 7(1), 29969 (2012)

[19] Kim, M.J., Chang, K.W., Park, S.H., Chang, W.S., Jung, H.H., Chang, J.W.: Stimulation-induced side effects of deep brain stimulation in the ventralis intermedius and posterior subthalamic area for essential tremor. Frontiers in Neurology 12, 678592 (2021)

[20] Vitek, J.L., Ashe, J., Delong, M.R., Kaneoke, Y.: Microstimulation of primate motor thalamus: somatotopic organization and differential distribution of evoked motor responses among subnuclei. Journal of neurophysiology 75(6), 2486–2495 (1996)

[21] Crowell, A.L., Ryapolova-Webb, E.S., Ostrem, J.L., Galifianakis, N.B., Shimamoto, S., Lim, D.A., Starr, P.A.: Oscillations in sensorimotor cortex in movement disorders: an electrocorticography study. Brain 135(2), 615–630 (2012)

[22] Gatev, P., Darbin, O., Wichmann, T.: Oscillations in the basal ganglia under normal conditions and in movement disorders. Movement disorders 21(10), 1566–1577 (2006)

[23] Kondylis, E.D., Randazzo, M.J., Alhourani, A., Lipski, W.J., Wozny, T.A., Pandya, Y., Ghuman, A.S., Turner, R.S., Crammond, D.J., Richardson, R.M.: Movement-related dynamics of cortical oscillations in parkinson’s disease and essential tremor. Brain 139(8), 2211–2223 (2016)

[24] Horn, A., Kühn, A.A.: Lead-dbs: a toolbox for deep brain stimulation electrode localizations and visualizations. Neuroimage 107, 127–135 (2015)

[25] Patriat, R., Palnitkar, T., Chandrasekaran, J., Sretavan, K., Braun, H., Yacoub, E., McGovern III, R.A., Aman, J., Cooper, S.E., Vitek, J.L., et al.: Dimani: diffusion mri for anatomical nuclei imaging—application for the direct visualization of thalamic subnuclei. Frontiers in Human Neuroscience 18, 1324710 (2024)

[26] Pelzer, E.A., Melzer, C., Timmermann, L., Cramon, D.Y., Tittgemeyer, M.: Basal ganglia and cerebellar interconnectivity within the human thalamus. Brain Structure and Function 222, 381–392 (2017)

[27] Karna, A., Gibert, K.: Automatic identification of the number of clusters in hierarchical clustering. Neural Computing and Applications 34(1), 119–134 (2022)

[28] Baker, M.R., Klassen, B.T., Jensen, M.A., Ojeda Valencia, G., Heydari, H., Ince, N.F., Müller, K.-R., Miller, K.J.: Parameterization of intraoperative human microelectrode recordings: Linking action potential morphology to brain anatomy. PLOS Computational Biology 21(6), 1013184 (2025)

[29] Herron, J., Kullmann, A., Denison, T., et al.: Challenges and opportunities of acquiring cortical recordings for chronic adaptive deep brain stimulation. Nature Biomedical Engineering 9, 606–617 (2025) 10.1038/s41551-024-01314-3

[30] Merk, T., Köhler, R.M., Brotons, T.M., et al.: Invasive neurophysiology and whole brain connectomics for neural decoding in patients with brain implants. Nature Biomedical Engineering (2025) 10.1038/s41551-025-01467-9

[31] Dixon, T.C., Strandquist, G., Zeng, A., et al.: Movement-responsive deep brain stimulation for parkinson’s disease using a remotely optimized neural decoder. Nature Biomedical Engineering (2025) 10.1038/s41551-025-01438-0

[32] Bouthour, W., Mgevand, P., Donoghue, J., Lüscher, C., Birbaumer, N., Krack, P.: Biomarkers for closed-loop deep brain stimulation in parkinson disease and beyond. Nature Reviews Neurology 15(6), 343–352 (2019)

[33] Bowyer, S.M.: Coherence a measure of the brain networks: past and present. Neuropsychiatric Electrophysiology 2, 1–12 (2016)

[34] Friston, K.J., Holmes, A.P., Worsley, K.J., Poline, J.-P., Frith, C.D., Frackowiak, R.S.: Statistical parametric maps in functional imaging: a general linear approach. Human brain mapping 2(4), 189–210 (1994)

[35] Cooley, J.W., Tukey, J.W.: An algorithm for the machine calculation of complex fourier series. Mathematics of computation 19(90), 297–301 (1965)

[36] Miller, K.J., Leuthardt, E.C., Schalk, G., Rao, R.P., Anderson, N.R., Moran, D.W., Miller, J.W., Ojemann, J.G.: Spectral changes in cortical surface potentials during motor movement. Journal of Neuroscience 27(9), 2424–2432 (2007)

[37] Miller, K.J., Honey, C.J., Hermes, D., Rao, R.P., Ojemann, J.G., et al.: Broadband changes in the cortical surface potential track activation of functionally diverse neuronal populations. Neuroimage 85, 711–720 (2014)

[38] Klassen, B.T., Baker, M.R., Jensen, M.A., Ojeda Valencia, G., Miller, K.J.: Spectral changes in motor thalamus field potentials during movement. Journal of neurophysiology 133(1), 101–108 (2025)

[39] Kingma, D.P., Ba, J.: Adam: A method for stochastic optimization. arXiv preprint arXiv:1412.6980 (2014)

[40] Klauschen, F., Dippel, J., Keyl, P., Jurmeister, P., Bockmayr, M., Mock, A., Buchstab, O., Alber, M., Ruff, L., Montavon, G., et al.: Toward explainable artificial intelligence for precision pathology. Annual Review of Pathology: Mechanisms of Disease 19(1), 541–570 (2024)

[41] Panaretos, V.M., Zemel, Y.: Statistical aspects of wasserstein distances. Annual review of statistics and its application 6, 405–431 (2019)

[42] Anders, C.J., Neumann, D., Samek, W., Müller, K.-R., Lapuschkin, S.: Software for dataset-wide xai: From local explanations to global insights with zennit, corelay, and virelay. PloS one 21(1), 0336683 (2026)

[43] Lapuschkin, S., Wäldchen, S., Binder, A., Montavon, G., Samek, W., Müller, K.-R.: Unmasking clever hans predictors and assessing what machines really learn. Nature communications 10(1), 1096 (2019)

[44] Yang, F., Zhang, Y., Wang, R., Li, Y., Huang, K., Yang, X.: Explainable artificial intelligence (xai) to find optimal in-silico biomarkers for cardiac drug toxicity evaluation. Scientific Reports 14(1), 6891 (2024) 10.1038/s41598-024-71169-w

[45] Sharma, G., Singh, P., Gupta, R.: An explainable ai-driven biomarker discovery framework for non-small cell lung cancer (nsclc). Computers in Biology and Medicine 155, 106612 (2023) 10.1016/j.compbiomed.2023.106612

[46] Pedregosa, F., Varoquaux, G., Gramfort, A., Michel, V., Thirion, B., Grisel, O., Blondel, M., Prettenhofer, P., Weiss, R., Dubourg, V., Vanderplas, J., Passos, A., Cournapeau, D., Brucher, M., Perrot, M., Duchesnay, É.: Scikit-learn: Machine learning in Python. Journal of Machine Learning Research 12, 2825–2830 (2011)

